# Asymptomatic carriers of the p.A53T SNCA mutation: data from the PPMI study

**DOI:** 10.1101/2021.12.04.21267242

**Authors:** Athina Maria Simitsi, Christos Koros, Maria Stamelou, Ion Beratis, Efthymia Efthymiopoulou, Dimitra Papadimitriou, Anastasia Bougea, Marina Picillo, Evangelia Stanitsa, Nikolaos Papagiannakis, Roubina Antonelou, Ioanna Pachi, Sokratis G. Papageorgiou, Paolo Barone, Leonidas Stefanis

## Abstract

**Introduction:** There has been great interest in the prodromal phase of Parkinson’s disease (PD), especially in subjects who are asymptomatic carriers of genetic mutations leading to PD because of the high risk to convert to PD. The objective of the present study was to assess non motor characteristics of asymptomatic p.A53T mutation carriers (A53T-AC) compared with healthy controls (HC).

**Methods:** We compared 12 A53T-AC with 36 matched HC enrolled into in the Parkinson’s Progression Markers Initiative (PPMI) study. Baseline data extracted from the PPMI database, contained demographics and non-motor symptoms (e.g. the Montreal Cognitive Assessment (MOCA) for cognition, the University of Pennsylvania Smell Identification Test (UPSIT) for olfaction, MDS-UPDRS I etc.)

**Results:** The mean UPSIT score was lower in A53T-AC vs HC (p =0.000). MoCA test showed a trend towards lower scores in A53T AC. We found a significant positive correlation between UPSIT score and MOCA in A53T-AC (r_s_ = 0,68, p=0,021) but not in HC. Total scores for MDS-UPDRS I did not differ between the groups but the subscore of anxiety was more prevalent in A53T-AC.

**Conclusion:** The more affected olfaction in A53T-AC may indicate that olfactory function is affected quite early in A53T carriers. The strong positive correlation between UPSIT and MOCA in the A53T-AC group may indicate that cognitive dysfunction and olfactory impairment progress alongside, prior to nigrostriatal degeneration. Anxiety was also more prevalent in A53T-AC and may represent an additional prodromal feature in this group of subjects.

## Introduction

SNCA, the gene encoding for the presynaptic protein alpha-synuclein, was the first gene linked to Parkinson’s disease (PD) [1]. The alpha-synuclein (AS) protein is the principal component of Lewy bodies (LBs) that are the neuropathological hallmark of the disease [2]. Variations at the SNCA locus are the major genetic risk factor for sporadic PD, while mutations cause familial disease. The discovery of the p.A53T SNCA mutation was a landmark in PD research [1]. Subsequently, a number of additional missense SNCA mutations and multiplications, inherited in an autosomal dominant fashion, have been described [3]. Patients with SNCA mutations are under intense scrutiny, given the manifest link of SNCA and AS to the pathophysiology, not only of familial, but also sporadic PD.

Recently there has been great interest in the investigation of the prodromal phase of PD, at a time before the onset of the motor symptoms of the disease. During this phase, a number of non-motor symptoms, such as depression, anxiety, constipation, sleep disturbances and olfactory deficits, may occur [4,5,6,7,8]. This prodromal phase of PD can be examined in large community cohorts of elderly subjects who are followed longitudinally, or in specific high-risk groups, such as those with idiopathic REM Sleep Behavior Disorder (RBD) or impaired olfaction [9.10]. Another group of special interest where cross-sectional or longitudinal studies can be performed, is that of subjects who are asymptomatic carriers of genetic mutations leading to PD. Such subjects have a high risk of converting to PD status, a risk that depends on the particular mutation and gene. The p.A53T SNCA mutation is highly penetrant, and asymptomatic carriers of this mutation offer a unique opportunity to examine the presyptomatic prodromal phase of the disease. Data from the literature concerning such asymptomatic carriers are scarce. Retrospective assessment in manifesting carriers of the mutation suggested that RBD and olfactory dysfunction could precede motor symptoms, but a rather basic assessment of 8 asymptomatic carriers at two time points within 2 years failed to reveal any distinctive features, especially since no comparative control group was assessed in parallel [11].

The objective of the present study was to assess baseline non motor characteristics of asymptomatic p.A53T mutation carriers (A53T-AC) compared with age - and sex - matched healthy controls (HC) enrolled in the Parkinson’s Progression Markers Initiative (PPMI) study.

## Materials and Methods

### Participants

Data used for this study were obtained from the PPMI database. The clinical trial identifier of the Parkinson’s Progression Markers Initiative (PPMI) study is NCT01141023. The study was approved by the ethics committee of both Attikon and Eginition Hospitals. Written informed consent was obtained from all patients under PPMI protocol, regardless of the enrolling site.

For our study we used the baseline dataset for both non-manifesting carriers of the p.A53T SNCA mutation and healthy controls from 33 participating outpatient Parkinson’s disease treatment centers worldwide.

Twelve asymptomatic p.A53T mutation carriers were recruited in the PPMI (ppmi-info.org) study. Nine of them (with a clinical follow up at the 1^st^ Department of Neurology, Eginition Hospital and 2nd Department of Neurology, Attikon Hospital, of the National and Kapodistrian University of Athens Medical School) were recruited from our center in Greece, one from the Beth Israel Medical Center, New York and two from the center at the University of Salerno, Italy. All asymptomatic carries enrolled in our PPMI site are first degree relatives of p.A53T PD patients who were screened for SNCA and glucocerebrosidase gene mutations (none harbored any GBA mutation). Because of the small number of asymptomatic carriers, and in order to increase the statistical power of the study, we recruited thirty-six healthy controls from the PPMI database, so that the ratio control to case would be equal to 3:1. We selected consecutive healthy controls from the PPMI database, using a proximity matching method, in order to obtain age- and sex-matched groups.

### Nonmotor symptoms assessment

For the assessment of non-motor symptoms the following scales were used: The Movement Disorder Society–sponsored Unified Parkinson’s Disease Rating Scale (MDS-UPDRS) I and I-Patient Questionnaire, the University of Pennsylvania Smell Identification Test (UPSIT) to assess olfaction, the REM sleep behavior disorder Questionnaire (RBDQ) in order to assess RBD, the Epworth Sleepiness Scale for daytime sleepiness(ESS), the Geriatric Depression Scale (GDS) to evaluate depression, Scales for Outcomes in Parkinson’s Disease–Autonomic questionnaire (SCOPA-AUT) scale for autonomic dysfunction. Compulsive behaviors were assessed using the Questionnaire for Impulsive-Compulsive Disorders in Parkinson’s Disease (QUIP). A detailed neuropsychological assessment was performed to evaluate cognitive function: The Montreal Cognitive Assessment (MoCA), the Hopkins Verbal Learning Test (HVLT), the Benton Judgement of Line Orientation test, the Letter Number Sequencing Test (LNST), and the Symbol Digit Modalities Test (SDMT) were administered. Semantic (animals, fruit, vegetables) and phonemic verbal fluency (words beginning with the Greek letter χ for the Greek participants) were assessed.

In addition, medications associated with the development of REM Sleep Behavior Disorder at the time of the evaluation were recorded. We also recorded whether the source of information for the RBDQ was from the participant, the caregiver or from both, because the information obtained by RBD questionnaires is insufficient especially when a bed partner is not available. The additional information of a caregiver is therefore valuable, especially if that person is also a bed partner.

### Statistical Analysis

For normally distributed scores, statistical significance was assessed with independent samples t test. For non-normally distributed scores, nonparametric Mann-Whitney U test was used. For dichotomous data analysis was performed using Chi–square test. A significance level of 0.05 was used in the demographic values. Regarding nonmotor symptoms, p < 0.0029 significance threshold was adopted to avoid type I errors (according to the Bonferroni method for multiple significance tests). For possible correlations-associations between numerical data we used the Spearman’s correlation coefficient for non-normally distributed scores. The statistical analyses were performed using commercially available software (SPSS version 20.0) (SPSS Inc., Chicago, IL).

## Results

### Demographic and Clinical Characteristics

The demographic features and clinical characteristics are summarized in the Table 1. Years of education were comparable (p = 0.923), as were medications associated with RBD (p=0.831) in the two groups. More specifically, two types of these medications were taken at the time of the study: antidepressants and beta-blockers. There was no statistically significant difference regarding the source of information for RBDQ (p=0.08), which was mainly provided only by the subjects themselves in both groups.

**Table 1.**
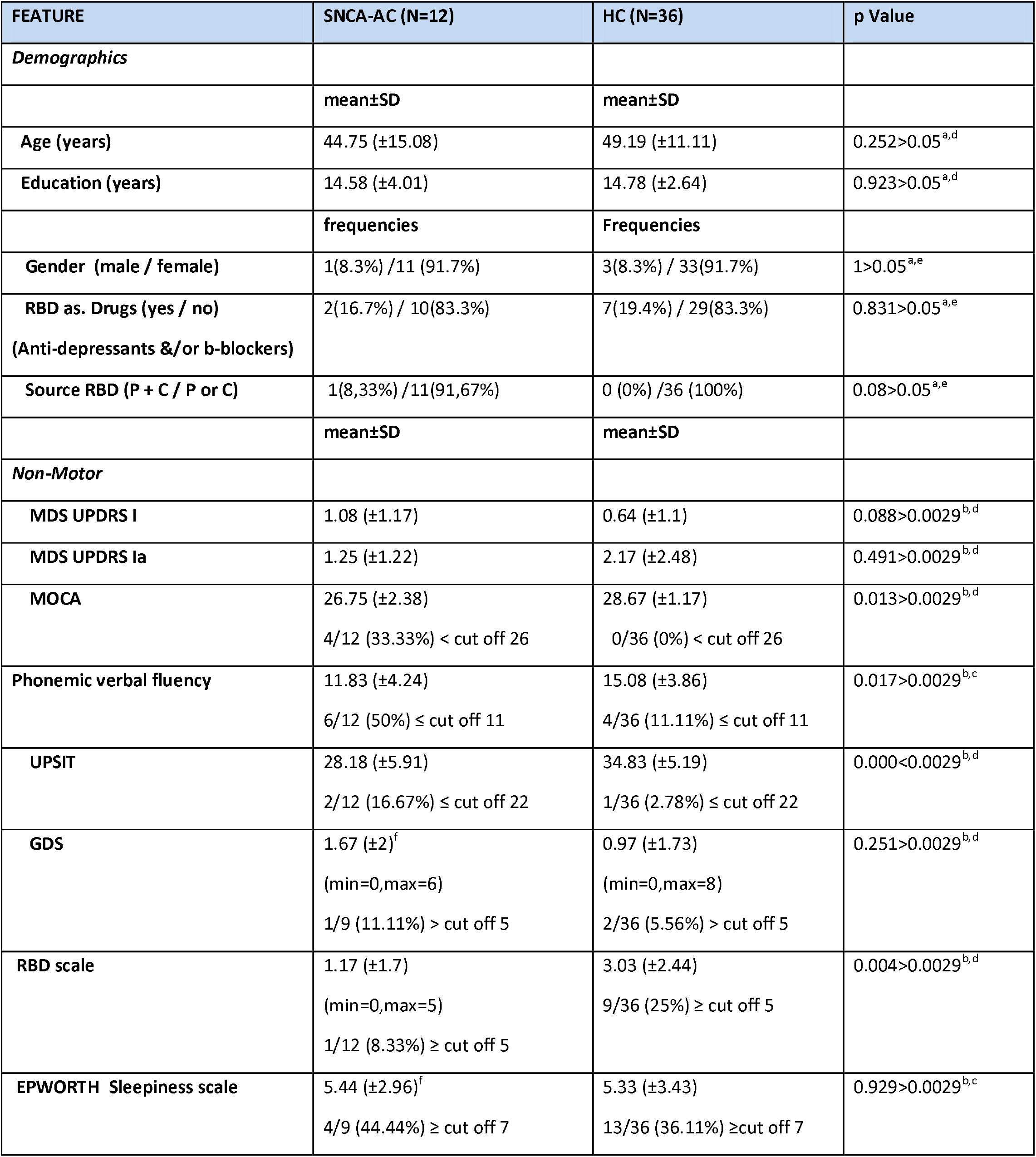
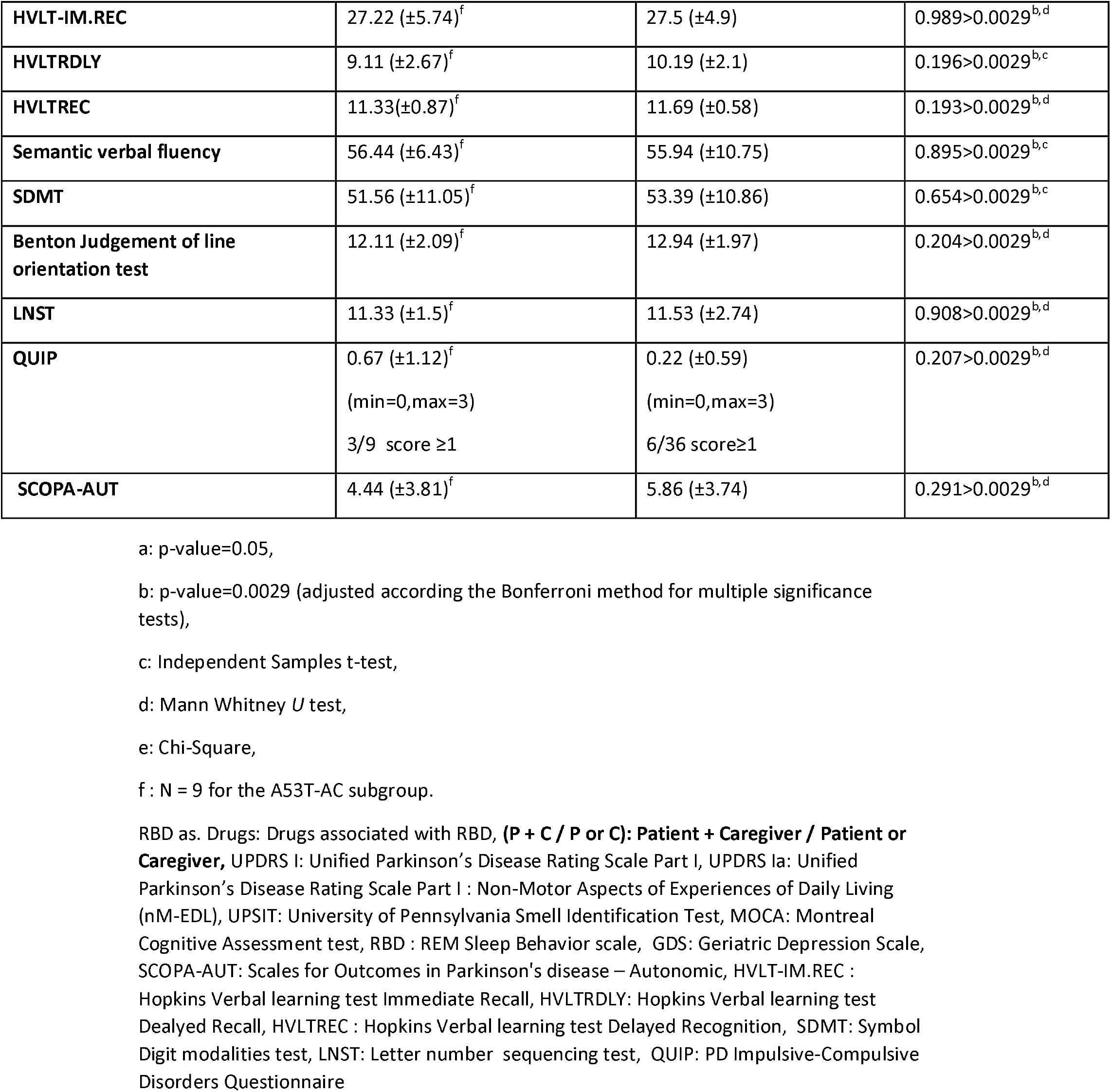
Demographics and non-motor characteristics in SNCA-AC vs. HC

### Nonmotor features

The mean UPSIT score was lower in A53T-AC vs HC (p =0.000). 16,67% of asymptomatic carriers had an abnormal UPSIT test (Table 2). As far as the neuropsychological assessment is concerned, MoCA test and phonemic verbal fluency showed a trend towards lower scores in A53T AC vs HC, but this trend did not retain statistical significance after Bonferroni correction for multiple comparisons (p=0,013 and p=0,017 respectively). All other neuropsychological tests did not differ between the two groups. We found a strong positive correlation, statistically significant between UPSIT score and MOCA in A53T-AC but not in HC (r_s_ = 0,68, p=0,021 and r_s_ = 0,054, p=0,756 respectively). SCOPA-AUT, QUIP and GDS scores were comparable. Regarding sleep-related problems, daytime sleepiness was not different between the groups. The mean RBDQ score was higher in HC vs AC (p=0.004) but did not survive Bonferroni correction.

**Table 2.** Unified Parkinson’s Disease Rating Scale PART I in SNCA-AC vs. HC

| FEATURE | SNCA-AC (N=12) | HC (N=36) | p Value | p1 Value |
| --- | --- | --- | --- | --- |
|  | mean±SD | mean±SD |  |  |
| *UPDRS I* |  |  |  |  |
| Cognitive impairment | 0 (±0) | 0.14 (±0.424)<br>(min=0,max=2) | 0.233>0.0083<sup>a,d</sup> | >0.0038<sup>c</sup> |
| Hallucinations and psychosis | 0 (±0) | 0 (±0) | 1>0.0083<sup>a,d</sup> | >0.0038<sup>c</sup> |
| Depressed mood | 0.17 (±0.577)<br>(min=0,max=2) | 0.11 (±0.319)<br>(min=0,max=1) | 0.857>0.0083<sup>a,d</sup> | >0.0038 |
| Anxious mood | 0.92 (±7.93)<br>(min=0,max=2) | 0.25 (±0.439)<br>(min=0,max=1) | 0.003<0.0083<sup>a,d</sup> | <0.0038 |
| Apathy | 0 (±0) | 0.08 (±0.28)<br>(min=0,max=1) | 0.307>0.0083<sup>a,d</sup> | >0.0038 |
| Features of DDS | 0 (±0) | 0.06 (±0.232)<br>(min=0,max=1) | 0.409>0.0083<sup>a,d</sup> | >0.0038 |
| *UPDRS Ia* |  |  |  |  |
| Sleep problems | 0.33 (±0.492)<br>(min=0,max=1) | 0.5 (±0.845)<br>(min=0,max=3) | 0.818>0.0071<sup>b,d</sup> | >0.0038 |
| Daytime sleepines | 0.17 (±0.389)<br>(min=0,max=1) | 0.47 (±696)<br>(min=0,max=2) | 0.179>0.0071<sup>b,d</sup> | >0.0038 |
| Pain and other sensations | 0 (±0) | 0.39 (±0.688)<br>(min=0,max=3) | 0.032>0.0071<sup>b,d</sup> | >0.0038 |
| Urinary problems | 0 (±0) | 0.11 (±0.319)<br>(min=0,max=1) | 0.233>0.0071<sup>b,d</sup> | >0.0038 |
| Constipation problems | 0.25 (±4.52)<br>(min=0,max=1) | 0.17 (±0.378)<br>(min=0,max=1) | 0.526>0.0071<sup>b,d</sup> | >0.0038 |
| Light headedness on standing | 0.25 (±0.622)<br>(min=0,max=2) | 0.11 (±0.319)<br>(min=0,max=1) | 0.561>0.0071<sup>b,d</sup> | >0.0038 |
| Fatigue | 0.25 (±0.452) | 0.42 (±0.692) | 0.537<0.0071<sup>b,d</sup> | >0.0038 |
|  | (min=0,max=1) | (min=0,max=3) |  |  |

- a: p-value=0.0083 (adjusted according the Bonferroni method for multiple significance tests) - b: p-value=0.0071 (adjusted according the Bonferroni method for multiple significance tests) - c: p-value=0.0038 (adjusted according the Bonferroni method for multiple significance tests) - d: Mann Whitney *U* test, UPDRS Part I: Unified Parkinson’s Disease Rating Scale Part I:Complex behaviors, UPDRS Part Ia: Unified Parkinson’s Disease Rating Scale Part I: Non-Motor Aspects of Experiences of Daily Living (nM-EDL).

Total scores for MDS-UPDRS I and I-Patient Questionnaire did not differ between the groups. As far as the subscores of the scales mentioned above, anxiety was more prevalent in A53T-AC (p=0.003 < p_MDS-UPDRSI-adj_=0.05/6=0.0083). Pain score was higher in HC but did not hold up after adjustment for multiple comparisons (p=0.032 > p_MDS-UPDRSIa- adj_=0.05/7=0.0071). The rest of the subscores were comparable. (See Table 2)

## Discussion

To our knowledge, this is the largest case series of asymptomatic p.A53T mutation carriers reported to date. One previous study from our group was longitudinal over a period of 2 years, included a smaller number of subjects, and entailed a rather rudimentary assessment, without a comparison group; this study identified only hints of non-motor symptoms in the asymptomatic cohort [11].

Our study shows that olfactory function is more affected in A53T-AC when compared to a group of matched HC. Hyposmia was present in 2/12 A53T-AC vs 1/36 in HC. This result, combined with our previous study in which hyposmia was more common in manifest A53T-PD compared to iPD[12], may indicate that olfactory function is affected quite early in A53T carriers, and thus is more impaired in this group, even when compared to iPD. This finding would be in agreement with one case of an asymptomatic A53T carrier, reported by Ricciardi et al [13], who demonstrated impaired olfaction, albeit in conjunction with a pathological DAT scan, while none of our cases had a dopaminergic deficit, as assessed with DAT SCAN per PPMI protocol. These findings are unlike those in other dominantly inherited genetic causes of PD, like GBA (N370S), where it seems that olfaction in AC is comparable to HC, while in AC-LRRK2 raw scores of UPSIT are comparable, but hyposmia is more prevalent[14,15].

Neuropsychological assessments revealed somewhat lower scores in global cognitive function, as assessed by the MoCA, and in verbal fluency in AC A53T compared to HC, but these findings did not survive after Bonferroni adjustment for multiple comparisons. We have previously shown that, compared to idiopathic PD, symptomatic carriers of the pA53T mutation are especially affected in cognitive functions related to a fronto-parietal network [12]. In the current work, other frontal lobe functions, as assessed by the LNST and the SDMT, and parietal functions, were not different between A53T-AC and HC. It remains plausible that A53T-AC manifest mild frontal lobe dysfunction that would be significant if a larger number of subjects were assessed. It is noteworthy that we and others have shown that cognitive impairment may occur in prodromal PD, based on a large community cohort; however, in that case, multiple cognitive domains were affected [16]. The present finding of a strong positive correlation between UPSIT and MOCA selectively in the A53T-AC group may indicate that subtle cognitive dysfunction evolves in parallel with olfactory impairment in prodromal A53T PD prior to nigrostriatal degeneration; however, the small sample size must be taken into account in this consideration, and further studies are warranted. These results are consistent with findings indicating that impaired olfaction is associated with prediction of cognitive dysfunction in iPD [17,18].

In our study paradoxically HC scored higher in RBDQ compared to A53T-AC. Given the poor predictive validity of such questionnaires for polysomnography-confirmed RBD [19,20], especially in the absence of confirmation of the information from a bed partner, as is the present study, and the paradoxical decrease of RBD features in A53T-AC compared to HC, we do not place too much weight on this difference, which, in any case, did not survive adjustment for multiple comparisons.

Comparing the subscores of UPDRS I, we also found anxiety to be more prevalent in A53T-AC. Depression and anxiety are common non motor features of PD that may precede motor symptoms [5]. Previous studies have shown that anxiety may be a prodromal feature, and it is included in the calculation of prodromal PD score[4]. A parkinsonian personality has also been described which is related to anxiety [21]. Thus, it is plausible that anxiety in our study may be a marker of the prodromal stage of the disease, reflecting possibly the involvement of extra-nigral brainstem structures. Alternatively, differences in scores could be driven by bias of both participants and investigators, who in most cases were aware of the participant’s genetic status. Awareness of a genetic status that predisposes heavily to a severe form of the disease, with a high socio-economic burden, that is already apparent in other family members, may put such subjects at risk for an anxiety disorder.

Limitations of this study include the small sample size, especially regarding male subjects, as our cohort included only one male subject. This may relate to the trend we have previously observed for female carriers of the mutation to manifest the disease less often and to have a milder phenotype [11]. In any case, our current study provides little information on asymptomatic male subjects with the p.A53T SNCA mutation. Another confounder is that, although the mean age of the A53T-AC subjects in this study was 45, some of them were very young (one of them was 28 and another 27 years old). Given the fact that the average age of onset of A53T-PD is around 46 years, it is quite possible that some of the subjects of the current study may have not yet entered the PD prodromal stage, while there is also the possibility that such asymptomatic subjects may not convert to PD during their lifetime, given that disease penetrance in carriers of this mutation is estimated at 80-90% [11]. However, it should be noted that all 7 asymptomatic carriers tested in the study of Wilson et al [22], including 5 from the present cohort, demonstrated a serotonergic deficit, indicating that the neurodegenerative process is under way. This serotonergic deficit may be related to the increased anxiety that we have observed. None of the subjects of the present study manifested a dopaminergic deficit, as assessed by DAT scan, thus they were all at the time of assessment at a stage of the disease prior to the onset of dopaminergic neurodegeneration. In the study of Wilson et al [22], premotor A53T carriers had no significant differences in non-motor symptoms compared with healthy controls, including UPSIT test. This discrepancy may be due to the different number of A53T-AC and HC in the two studies (only 5 subjects A53T-AC out of 12 of our study, were common in both studies) and UPSIT tests were made at different times and in different conditions (in our study the results of UPSIT were from the PPMI database).

Further studies, in particular longitudinal, are needed in asymptomatic A53T carriers, in order to determine the sequence of events associated with the early stages of neurodegeneration, and more particularly to assess whether olfactory function or other nonmotor symptoms can be used as a marker of the premotor stage of p.A53T-associated PD, the prototypical genetic synucleinopathy.

## Data Availability

All data produced in the present study are available upon reasonable request to the authors

## Acknowledgment

We are grateful to asymptomatic carriers of the p.A53T SNCA mutation for their participation in this project. We thank volunteer healthy controls who contributed samples for this study. This work was supported by the Michael J. Fox Foundation for Parkinson’s Research (https://www.ppmi-info.org/data).

## Authors’ Role

1. Research study: A. Conception and design / acquisition of data, B. Analysis and interpretation of data.
2. Manuscript: Drafting the article or revising it critically for important intellectual content
3. Final approval of the version to be submitted.

| **Name** | **Role** |
| --- | --- |
| **Athina Maria Simitsi** | 1,2,3 |
| **Christos Koros** | 2,3 |
| **Maria Stamelou** | 2,3 |
| **Ion Beratis** | 2,3 |
| **Efthymia Efthymiopoulou** | 2,3 |
| **Dimitra Papadimitriou** | 2,3 |
| **Anastasia Bougea** | 2,3 |
| **Marina Picillo** | 2,3 |
| **Evangelia Stanitsa** | 2,3 |
| **Nikolaos Papagiannakis** | 2,3 |
| **Roubina Antonelou** | 2,3 |
| **Ioanna Pachi** | 2,3 |
| **Sokratis G Papageorgiou** | 2,3 |
| **Paolo Barone** | 2,3 |
| **Leonidas Stefanis** | 1,2,3 |

## References

[1] Polymeropoulos MH, Lavedan C, Leroy E, Ide SE, Dehejia A, Dutra A, Pike B, Root H, Rubenstein J, Boyer R, Stenroos ES, Chandrasekharappa S, Athanassiadou A, Papapetropoulos T, Johnson WG, Lazzarini AM, Duvoisin RC, Di Iorio G, Golbe LI, Nussbaum RL, Mutation in the alphasynuclein gene identified in families with Parkinson’s disease, Science. 276 (5321) (1997) 2045–7.

[2] Spillantini MG, Schmidt ML, Lee VM, Trojanowski JQ, Jakes R, Goedert M, Alpha-synuclein in Lewy bodies, Nature. 388 (6645) (1997) 839–40.

[3] Koros C, Simitsi A, Stefanis L, Genetics of Parkinson’s Disease: Genotype-Phenotype Correlations, Int Rev Neurobiol. 132 (2017) 197–231.

[4] Bower JH, Grossardt BR, Maraganore DM, Ahlskog JE, Colligan RC, Geda YE, Therneau TM, Rocca WA, Anxious personality predicts an increased risk of Parkinson’s disease, Mov Disord. 25 (13) (2010) 2105–13

[5] Leentjens AF, Van den Akker M, Metsemakers JF, Lousberg R, Verhey FR, Higher incidence of depression preceding the onset of Parkinson’s disease: a register study, Mov Disord. 18 (4) (2003) 414–8.

[6] Iranzo A, Molinuevo JL, Santamaría J, Serradell M, Martí MJ, Valldeoriola F, Tolosa E, Rapid-eye-movement sleep behaviour disorder as an early marker for a neurodegenerative disorder: a descriptive study, Lancet Neurol. (7) (2006) 572–7.

[7] Mahlknecht P, Iranzo A, Högl B, Frauscher B, Müller C, Santamaría J, Tolosa E, Serradell M, Mitterling T, Gschliesser V, Goebel G, Brugger F, Scherfler C, Poewe W, Seppi K; Sleep Innsbruck Barcelona Group, Olfactory dysfunction predicts early transition to a Lewy body disease in idiopathic RBD, Neurology. 84(7) (2015) 654–8.

[8] Lin CH, Lin JW, Liu YC, Chang CH, Wu RM, Risk of Parkinson’s disease following severe constipation: a nationwide population-based cohort study, Parkinsonism Relat Disord. 20 (12) (2014) 1371–5.

[9] Chen H, Shrestha S, Huang X, Jain S, Guo X, Tranah GJ, Garcia ME, Satterfield S, Phillips C, Harris TB; Health ABC Study, Olfaction and incident Parkinson disease in US white and black older adults, Neurology. 89 (14) (2017) 1441–1447.

[10] Rees RN, Noyce AJ, Schrag A, The prodromes of Parkinson’s disease, Eur J Neurosci. 49 (3) (2019) 320–327.

[11] Papadimitriou D, Antonelou R, Miligkos M, Maniati M, Papagiannakis N, Bostantjopoulou S, Leonardos A, Koros C, Simitsi A, Papageorgiou SG, Kapaki E, Alcalay RN, Papadimitriou A, Athanassiadou A, Stamelou M, Stefanis L, Motor and Nonmotor Features of Carriers of the p.A53T Alpha-Synuclein Mutation: A Longitudinal Study Mov Disord. 31(8) (2016) 1226–30.

[12] Koros C, Stamelou M, Simitsi A, Beratis I, Papadimitriou D, Papagiannakis N, Fragkiadaki S, Kontaxopoulou D, Papageorgiou SG, Stefanis L, Selective cognitive impairment and hyposmia in p.A53T SNCA PD vs typical PD, Neurology. 90 (10) (2018) e864–e869

[13] Ricciardi L, Petrucci S, Di Giuda D, Serra L, Spanò B, Sensi M, Ginevrino M, Cocciolillo F, Bozzali M, Valente EM, Fasano A, The Contursi Family 20 Years Later: Intrafamilial Phenotypic Variability of the SNCA p.A53T Mutation, Mov Disord. 31 (2) (2016) 257–8.

[14] Simuni T, Uribe L, Cho HR, Caspell-Garcia C, Coffey CS, Siderowf A, Trojanowski JQ, Shaw LM, Seibyl J, Singleton A, Toga AW, Galasko D, Foroud T, Tosun D, Poston K, Weintraub D, Mollenhauer B, Tanner CM, Kieburtz K, Chahine LM, Reimer A, Hutten SJ, Bressman S, Marek K; PPMI Investigators, Clinical and dopamine transporter imaging characteristics of non-manifest LRRK2 and GBA mutation carriers in the Parkinson’s Progression Markers Initiative (PPMI): a cross-sectional study, Lancet Neurol. 19 (1) (2020) 71–80.

[15] Sierra M, Sánchez-Juan P, Martínez-Rodríguez MI, González-Aramburu I, García-Gorostiaga I, Quirce MR, Palacio E, Carril JM, Berciano J, Combarros O, Infante J, Olfaction and imaging biomarkers in premotor LRRK2 G2019S-associated Parkinson disease, Neurology. 80 (7) (2013) 621–6.

[16] Bougea A, Maraki MI, Yannakoulia M, Stamelou M, Xiromerisiou G, Kosmidis MH, Ntanasi E, Dardiotis E, Hadjigeorgiou GM, Sakka P, Anastasiou CA, Stefanis L, Scarmeas N, Higher probability of prodromal Parkinson disease is related to lower cognitive performance, Neurology. 92 (19) (2019) e2261–e2272.

[17] Kang SH, Lee HM, Seo WK, Kim JH, Koh SB, The combined effect of REM sleep behavior disorder and hyposmia on cognition and motor phenotype in Parkinson’s disease, J Neurol Sci. 368 (2016) 374–8.

[18] Fullard ME, Tran B, Xie SX, Toledo JB, Scordia C, Linder C, Purri R, Weintraub D, Duda JE, Chahine LM, Morley JF, Olfactory impairment predicts cognitive decline in early Parkinson’s disease, Parkinsonism Relat Disord. 25 (2016) 45–51.

[19] Li K, Li SH, Su W, Chen HB, Diagnostic accuracy of REM sleep behaviour disorder screening questionnaire: a meta-analysis, Neurol Sci. 38 (6) (2017) 1039–1046.

[20] Halsband C, Zapf A, Sixel-Döring F, Trenkwalder C, Mollenhauer B, The REM Sleep Behavior Disorder Screening Questionnaire is not Valid in De Novo Parkinson’s Disease, Mov Disord Clin Pract. 5 (2) (2018) 171–176.

[21] Ishihara L, Brayne C. What is the evidence for a premorbid parkinsonian personality: a systematic review, Mov Disord. 21 (8) (2006) 1066–72.

[22] Wilson H, Dervenoulas G, Pagano G, Koros C, Yousaf T, Picillo M, Polychronis S, Simitsi A, Giordano B, Chappell Z, Corcoran B, Stamelou M, Gunn RN, Pellecchia MT, Rabiner EA, Barone P, Stefanis L, Politis M, Serotonergic pathology and disease burden in the premotor and motor phase of A53T α-synuclein parkinsonism: a cross-sectional study, Lancet Neurol. 18 (8) (2019) 748–759.

